# Quantifying individual-level heterogeneity in infectiousness and susceptibility through household studies

**DOI:** 10.1101/2022.12.02.22281853

**Authors:** Thayer L Anderson, Anjalika Nande, Carter Merenstein, Brinkley Raynor, Anisha Oommen, Brendan J Kelly, Michael Z Levy, Alison L Hill

## Abstract

The spread of SARS-CoV-2, like that of many other pathogens, is governed by heterogeneity. “Superspreading,” or “over-dispersion,” is an important factor in transmission, yet it is hard to quantify. Estimates from contact tracing data are prone to potential biases due to the increased likelihood of detecting large clusters of cases, and may reflect variation in contact behavior more than biological heterogeneity. In contrast, the average number of secondary infections per contact is routinely estimated from household surveys, and these studies can minimize biases by testing all members of a household. However, the models used to analyze household transmission data typically assume that infectiousness and susceptibility are the same for all individuals or vary only with predetermined traits such as age. Here we develop and apply a combined forward simulation and inference method to quantify the degree of inter-individual variation in both infectiousness and susceptibility from observations of the distribution of infections in household surveys. First, analyzing simulated data, we show our method can reliably ascertain the presence, type, and amount of these heterogeneities with data from a sufficiently large sample of households. We then analyze a collection of household studies of COVID-19 from diverse settings around the world, and find strong evidence for large heterogeneity in both the infectiousness and susceptibility of individuals. Our results also provide a framework to improve the design of studies to evaluate household interventions in the presence of realistic heterogeneity between individuals.

## Introduction

In the early months of the COVID-19 pandemic, contact tracing efforts revealed that superspreading events — when a single infected individual transmits to a large number of secondary individuals — play an important role in the spread of SARS-CoV-2 [1–6]. According to a systematic review, 21/26 studies quantifying SARS-CoV-2 superspreading reported significant over-dispersion in transmission and found that between 1% to 39% of individuals were responsible for 80% of secondary infections (with estimates for the negative binomial dispersion parameter k ranging from 0.01 to 0.72) [7]. Superspreading has also been recognized as a key feature of transmission dynamics for a wide variety of other infections including SARS [8,9], MERS [10], smallpox [8], ebola [11], tuberculosis [12–14], and HIV [15]. These dynamics highlight the limitations of summarizing transmission patterns using only average quantities, like the basic reproductive number, R_0_ [16]. More generally, individuals may vary in both their infectiousness (propensity for onward transmission if infected) and susceptibility (propensity to become infected given exposure). This heterogeneity is caused by a mix of of host behavior, host-pathogen interactions within the body, and environmental factors [8, 11, 17–35]. For example, variation in infectiousness can stem from heterogeneity in the number, type, and intensity of an index individual’s contacts; [27]; heterogeneity in pathogen shedding in respiratory, genital or fecal excretions [28–34]; infectious period duration [8, 11, 35], and pathogen survival outside the host body [36,36–39]. Host susceptibility has been observed to vary based on immune status (due to genetics [17], age [23], disease (e.g., AIDS [40]), medication (e.g., [24, 25]), temperature (e.g., [41]), or previous exposure due to infection or vaccination [26]) and other phenotypes (e.g., [18–22]). Models have demonstrated that variation in infectiousness plays a crucial role in determining the extinction probability of an outbreak following introduction into a new population [8], the final size of an epidemic [16], the efficacy of contact tracing [42], the rate of emergence of pathogen variants [43], and the optimal allocation of measures to prevent infection or reduce transmission [44]. Variation in host susceptibility affects the epidemic growth rate, the herd immunity threshold and the final epidemic size [26, 45, 46], and provides opportunities for targeting preventative measures such as vaccination [44, 47–52].

While characterizing variation in infectiousness and susceptibility is a critical part of designing accurate mathematical models and effective public health interventions, it is difficult to quantify in practice. Population-level epidemic growth patterns that are used to infer R_0_ (e.g., [53, 54]) can rarely uniquely identify variation among individuals. Contact tracing studies are the most common tool used to estimate the degree of variation in infectiousness, but they suffer from possible biases. Chains of infection may be underestimated when relying on individuals naming known contacts, but at the same time, superspreading events may be more likely to be detected by surveillance efforts than “normal” chains of infection due to the number of individuals they involve [7, 55]. Lloyd-Smith found that joint estimation of R_0_ and the degree of variation in infectiousness was particularly difficult with certain types of censoring present in outbreak data [56]. Variation caused by host behavior — like travel patterns — as opposed to biological or environmental causes might have an outsized effect on these studies. Public policy measures such as mask mandates can increase the fraction of transmission stemming from superspreading events, as those events are likely to involve greater risk and fewer precautions [55]. Mild cases of disease can involve lower viral load and therefore lower probability of onwards transmission but also a lower probability of detection, which may further bias estimates of heterogeneity (e.g. [57]).

Household transmission studies are a common tool in infectious disease epidemiology that sidestep many of these possible biases. Households are a context where it is known – or can be reasonably assumed – that all individuals are in close contact with each other and where infection status can be ascertained for the entire group. However, household studies typically only focus on calculating the *average* probability of infection per contact, known as the secondary attack risk (SAR). Simple estimates of the SAR assume all infected secondary contacts are caused directly by the primary case (reviewed in [58], examples in [59, 60]), while other studies account for multiple generations of infections using the ‘Reed-Frost’ or ‘chain binomial’ model [61]. When household studies evaluate heterogeneity at all, it is commonly by stratifying cases by a pre-specified individual trait (like host age) [62]. Reed-Frost models have been extended to “multitype” versions in which distinct classes of individuals have different infectivities [63, 64] and “collective” versions where infectiousness varies continuously across individuals [63,65,66], and have been used to estimate jointly the SAR and variation in infectiousness for the 1918 influenza pandemic [67], 2009 “swine flu” (H1N1) [68], and COVID-19 [69, 70]. A stochastic, discrete-generation model chain binomial has the benefit of admitting analytic solutions for the outbreak size distribution in a finite, well-mixed population like a household, and is often mathematically equivalent to continuous time models [63, 71]. However, its use requires the absence of any parameter dependence on calendar time [63] and is therefore unsuitable in circumstances where interventions such as availability of vaccines/therapeutics, out-of-home isolation, or changes in household composition play a substantial role in infection dynamics [63] (e.g. [72]).

In this study we develop a method to simultaneously estimate the SAR and the degree of heterogeneity in infectiousness and susceptibility from household transmission studies, using arbitrary models of disease dynamics and arbitrary distributions of individual-level traits. Our approach combines direct model simulation with formal likelihood-based parameter inference. We can consistently estimate key epidemiological parameters with modest computational resources, with the precision of parameter estimates and the power to identify the impact of interventions increasing with both study size and the inclusion of households with more than two individuals. We find that ignoring heterogeneities, when they exist, can lead to biased estimates of the SAR. Applying this method to multiple existing households studies of COVID-19 in diverse settings, we quantify both types of transmission heterogeneity. We hypothesize that studying the variability of pathogen spread within households can limit the sources of measured heterogeneity to those sources that are tied most closely to host-pathogen interactions and biology.

## Methods

### Model of transmission within households

The focus of our model is a population in which individuals are divided up into households of different sizes (Figure 1C). Each household is assumed to be a well-mixed population with a single introduction of infection. We consider diseases with finite infectious periods (e.g., diseases that lead to eventual recovery and immunity) and assume the outbreak within each household is complete by the end of the study period. These assumptions are valid when the study period is long relative to the infectious period, and when risk of transmission from the community is much smaller than the risk of transmission from within a household. Our direct simulation method allows for relaxation of these assumptions if desired. We track the number of individuals within each household who were ever infected (final epidemic size).

**Figure 1:**
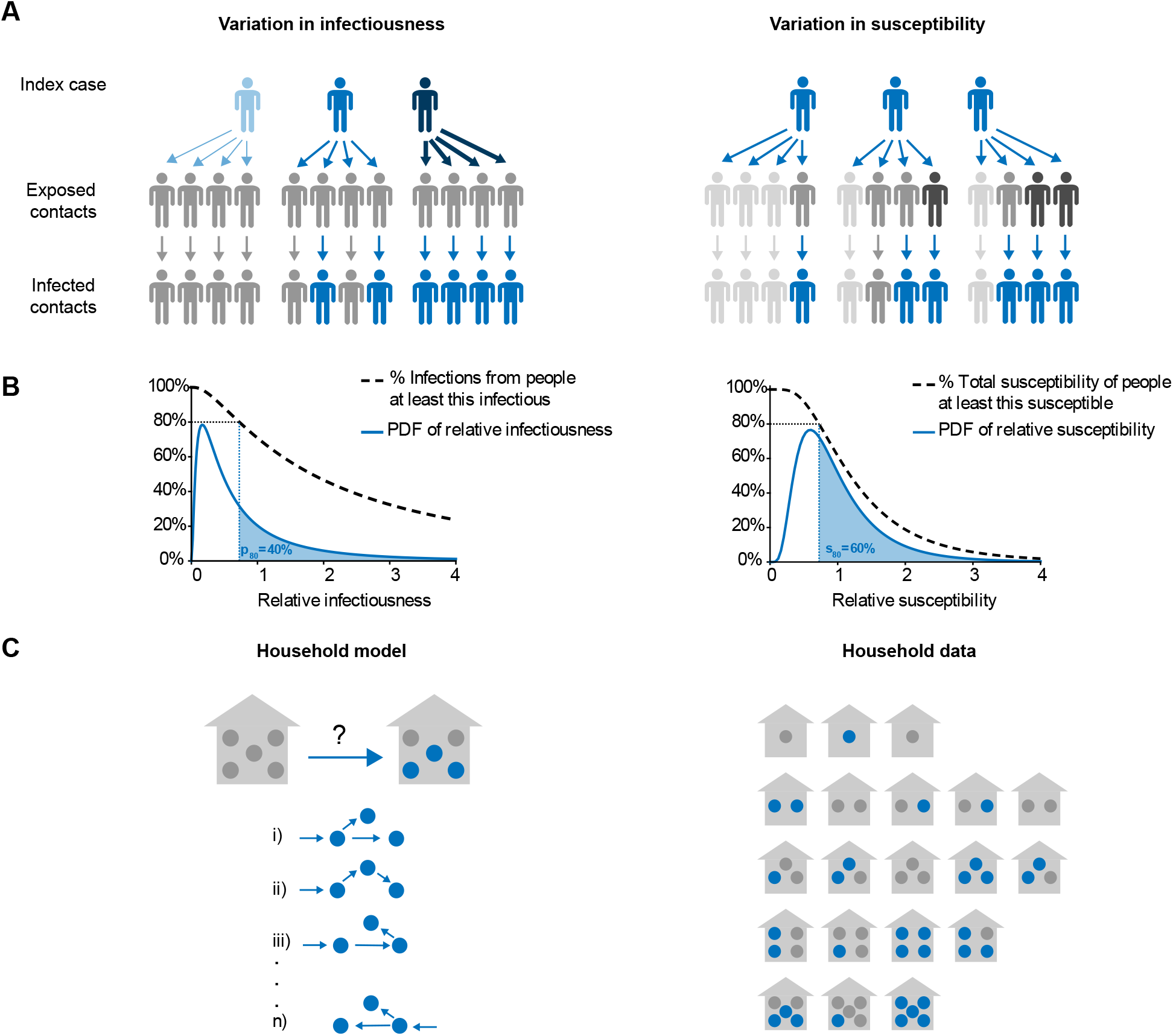
Model schematic for individual-level heterogeneity and household transmission studies. A) The effect of variation in infectiousness (left) and susceptibility (right) on an imagined chain of infections across two generations of spread. Grey = uninfected, Blue = infected. Darker color = more susceptible/infectious. B) Left: Probability density function for relative individual infectiousness and representation of relationship between p_80_ (shaded area), relative infectiousness (x-axis), and the fraction of infections from people at least that infectious (y-axis). Right: Probability density for relative susceptibility, and representation of relationship between s_80_ (shaded area), relative susceptibility (x-axis), and fraction of total susceptibility held by people at least that susceptible (y-axis). C) Left: Possible infection chains, simulated by the model, that give rise to the final observed outbreak size in a household. Right: Example dataset comprised of the distribution of household outbreak sizes.

When an infected individual and a susceptible individual are in the same household, the average probability per unit time that infection passes between them is β, but the specific rate varies between pairs of individuals due to the individual-level heterogeneity in per-contact infectiousness and per-contact susceptibility. We summarize the transmission potential of an infection with the household secondary attack risk (SAR), defined as the average probability of infection per household contact caused by a single initial introduction of the pathogen (i.e., not taking into account chains of infection beyond the initial secondary).The mathematical relationship between the SAR and β depends on the nature of the variation in infectiousness and susceptibility across individuals, as well as on the distribution of the duration of infectious periods (see Supplementary Methods).

Our inference framework allows for the specification of any model of transmission within households. For simplicity and for consistency with standard methods for the simulation of COVID-19 dynamics, we choose to model transmission with a stochastic SEIR (susceptible, exposed, infectious, recovered) process. We assume that the times spent in the exposed and infectious states follow lognormal distributions, a long-tailed distribution chosen because it simplifies the calculation of expected secondary infections (see Supplemental Methods). For the latent period, we choose parameters of the distribution such that the average length is 3.5± 2.5 days, and for the infectious period, we choose them so that the length is 6± 2.5 days (consistent with Zhao et al. [73] and references therein).

### Modeling heterogeneity in infectiousness and susceptibility

Under our model, each individual in a household is assigned two traits: one that determines their relative per-contact infectiousness and one that determines their relative per-contact susceptibility (Figure 1). In the population, these traits are continuously-valued random variables. Beyond these traits, individuals are identical and we assume the distributions of traits are independent of factors such as age or biological sex.

The most important consequence of heterogeneity in infectiousness is over-dispersion in the expected number of secondary infections caused by each individual. As a result, it is common to quantify the heterogeneity in infectiousness in terms of the distribution of expected infections. Following Lloyd-Smith et al., it is common to model the distribution as a negative binomial distribution with “dispersion parameter” k [8]. We take an alternative approach [15, 74] and quantify heterogeneity with p_80_, defined as the (smallest) fraction of individuals responsible for 80% of secondary infections on average (Figure 1, Supplemental Methods). When p_80_ = 80%, each individual is equally infectious. For p_80_ < 80%, a smaller fraction of individuals is responsible for a greater portion of secondary infections (i.e. the distribution is over-dispersed [55, 74]). We model the distribution of relative infectiousness in the population as a lognormal random variable with natural-scale mean of 1, and we numerically solve for the variance given a desired value of p_80_.

We describe variation in relative susceptibility through the quantity s_80_, the smallest fraction of the population that must be infected in order to reduce the total remaining susceptibility by 80% of its initial total. This approach is based on the fact that as an infection spreads through a population with heterogeneous susceptibilities, the average susceptibility in the uninfected population decreases over time as more susceptible individuals are infected first with greater probability. When s_80_ = 80%, all individuals are equally susceptible. If s_80_ = 20%, then the most susceptible 20% of individuals are responsible for 80% of the total susceptibility.

If they were all infected, the average susceptibility of uninfected individuals would be 20% of its initial mean. We model susceptibility variation as another lognormal random variable with mean 1, and we calculate the variance based on the specified s_80_. When infection is first introduced into a household, the index individual is randomly assigned with probability in proportion to the susceptibility of each household member.

### Parameter inference

To make inferences, we calculate the likelihood of observing a particular dataset under the proposed model of household transmission for each possible set of model parameters (θ), and then find the set of parameters that maximizes this likelihood function. The relevant data consists of the number of infections in each household in a cohort, where households can be grouped by number of members (Figure 1C). Formally, the data consists of a set of y_*n,k*_, the total number of households of size n observed to have k infections.

We calculate the probability p_*M*_ (n, k, θ) under our model M that a single household of size n observes L k infections after a single introduction by simulating a large volume of households of size n forward in time from their initial state and treating the frequency of occurrences for the different k’s as the probability of observing that many infections. Then, the likelihood of observing the full dataset given the model, (Y, θ), is given by the multinomial:

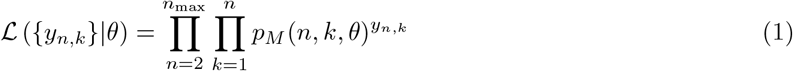

By varying the model parameters θ, we construct a likelihood surface. The maximum likelihood estimate of the parameter values (MLE) is the position of θ that produces the greatest likelihood out of all tested combinations of parameter values. The likelihood surface can be viewed as a (discrete) probability density function of a posterior probability distribution in a Bayesian framework by making the assumption of uniform priors on all parameters and then normalizing. To calculate 95% confidence intervals (equivalently, credible intervals), we rank each tested θ value set from most probable to least probable, include θ values in order of descending probability until at least 95% of the overall probability is represented, and then determine the extreme values for each parameter.

In this paper the unknown parameters that we infer are (1) p_80_, (2) s_80_, and (3) the average transmission rate β — although we actually infer a transformation of β, SAR, the average secondary attack risk in the population (Supplemental Methods). For inference on real data from COVID-19 outbreaks and for benchmarking tests on simulated data, we assumed that the other model parameters – the duration of the latent period and infectious period – are known exactly (i.e. not inferred).

## Results

### The effect of heterogeneity on household outbreaks

Simulating the spread of infection in households, we found that the average risk to a susceptible contact (SAR), the amount of inter-individual variation in infectiousness (p_80_), and the amount of inter-individual variation in susceptibility (s_80_) each affect the final size of household outbreaks in distinct ways (Figure Predictably, we observed that a greater SAR increases the frequency with which an outbreak caused by a single introduction will spread to other individuals in a household and consequently lowers the within-household extinction probability (i.e., the frequency of outbreaks of size 1) and increases the frequency of larger outbreaks (Figure 2A).

**Figure 2:**
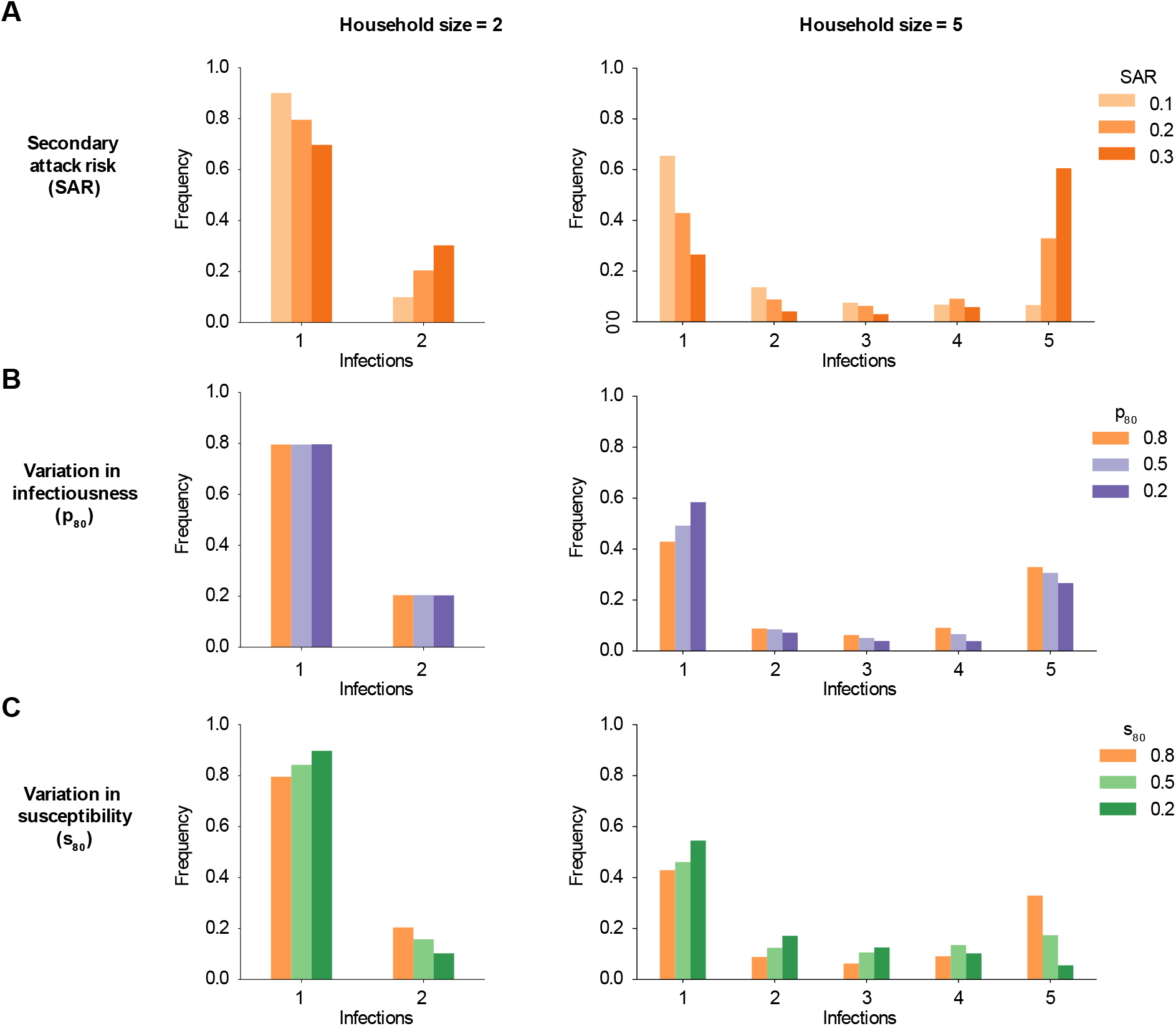
The effect of heterogeneity in infectiousness and susceptibility on the distribution of infections in a household outbreak. Histograms of simulated infection outcomes (including index case) in households of different sizes after a single introduction of a pathogen. x-axis: number of infections in household. y-axis: frequency that that number of infections is observed in all households with at least one infection. (A): Comparison of infections in households of size 2 (left column) and size 5 (right column) for different values of the average secondary attack risk (SAR). (B) Simulated infections with SAR = 20% and different amounts of infectiousness variation (more when p_80_ is smaller: darker purple). (C): Simulated infections for SAR = 20% and different amounts of susceptibility variation (more when s_80_ is smaller: darker green).

Variation in infectiousness (more superspreading) increases “feast or famine” dynamics: outbreaks are more likely to go extinct, and more likely to infect all household members provided that they do not go extinct (Figure 2B). Susceptibility variation, in contrast, increases the probability of extinction but also concentrates the distribution of final sizes at intermediate numbers of infections and reduces the probability that the entire household is infected (Figure 2C). Since a more susceptible individual is proportionally more likely to be the site of introduction, other household members have on average lower susceptibility and a reduced risk of secondary infection. Susceptibility variation therefore impacts the outbreak size even in households of size 2 for a fixed average infection probability (SAR).

As an example, in a household of size 5 with SAR = 20% and no variation in infectiousness or susceptibility, an outbreak results in a single infection 43% of the time, 3 infections 6% of the time, and 5 infections 33% of the time. In the same household when the most infectious 20% of individuals are responsible for 80% of secondary infections (p_80_ = 20%), the frequency of outbreaks of size 1, 3, and 5 becomes 58%, 4%, and 27%. Alternatively, if there is variation in susceptibility such that the most susceptible 20% of individuals hold 80% of the population’s total susceptibility (s_80_ = 20%), the frequency of outbreaks of size 1, 3, and 5 becomes 55%, 13%, and 5%. These distinct patterns in the final size distribution of household outbreaks suggest that it is possible to infer the SAR and the heterogeneities simultaneously from data.

### Inferring heterogeneity parameters from household outbreak sizes

Before trying to infer heterogeneity from real-world data collected in studies of household outbreaks, we first confirmed that our inference method could recover the input parameters used to generate simulated data. We evaluated scenarios where only one kind of heterogeneity was present, as well as scenarios where there was heterogeneity in both traits (Figure 3). We found that the SAR can be accurately and precisely inferred regardless of the kinds and amount of heterogeneity present, provided that the model is correctly specified. For simulated datasets of 5,000 individuals with high heterogeneity in both traits, the maximum likelihood estimate (MLE) of the SAR was within 5% points of the true value (20%) 95% of the time. For any individual simulated dataset with 5,000 individuals, the uncertainty in the estimated SAR was around ± 7% points. However, when the model used for inference doesn’t incorporate heterogeneity while it is present in the data-generating dynamics, significant bias is introduced into the estimate of the SAR — particularly when a high degree of susceptibility variation is present. For the simulated datasets with SAR = 20% described above, the MLE estimates with a misspecified model were centered on ≈ 19% in the absence of susceptibilty variation and ≈ 11% in its presence).

**Figure 3:**
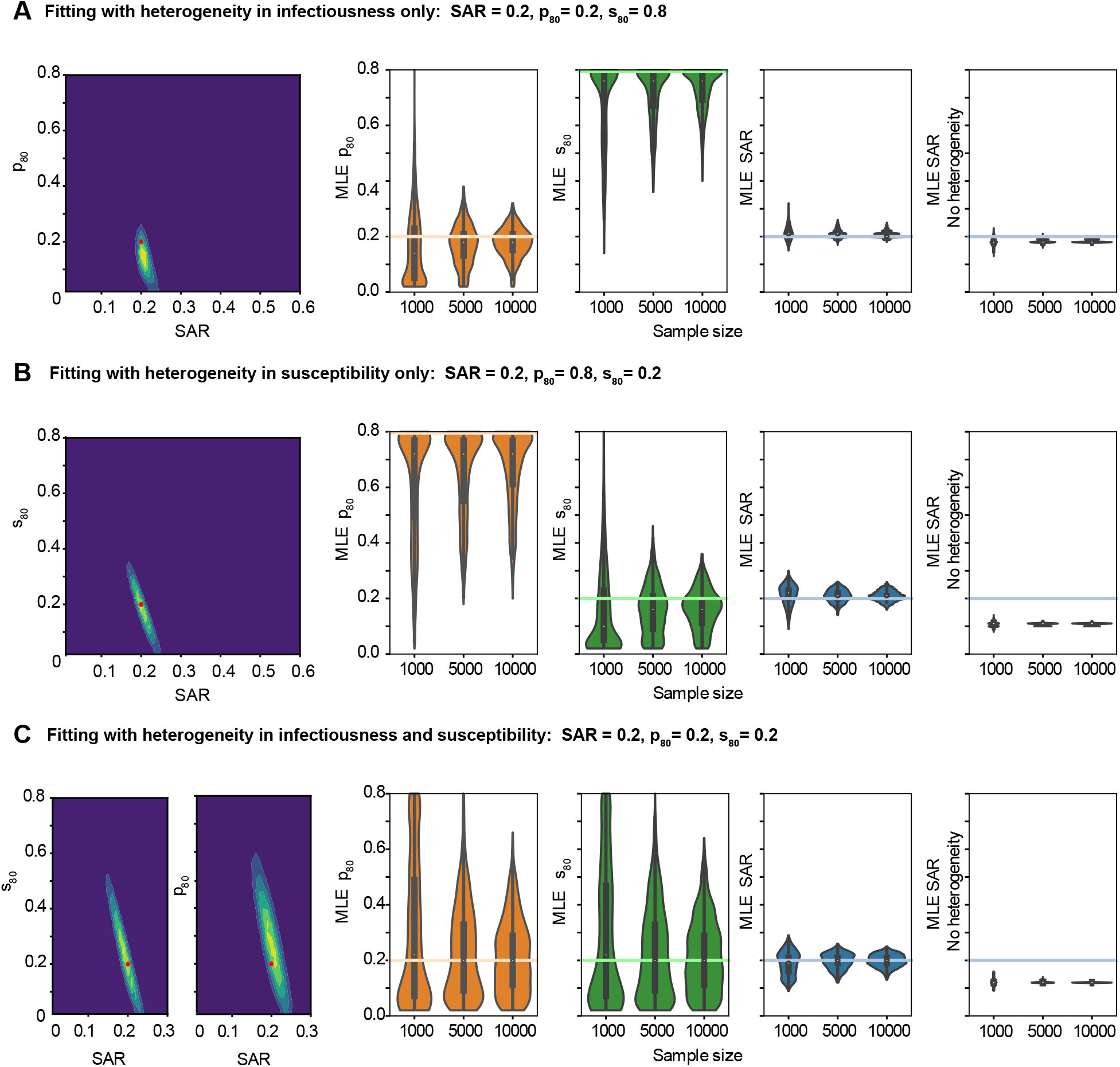
Maximum likelihood estimates for SAR and degree of heterogeneity on simulated data in a variety of parameter regimes. Simulated datasets were generated with 1,000, 5,000, or 10,000 individuals divided among households with a household size distribution taken from the United States census. A) Simulated data includes only variation in infectiousness (p_80_). B) Simulated data includes only variation in susceptibility (s_80_). C) Simulated data includes variation in both infectiousness and susceptibility. For (A)-(C), the left panel shows the likelihood surface describing the probability of the model parameters given a simulated dataset (yellow is more likely, blue is less likely). This two dimensional surface marginalizes over the third, not shown, parameter. The red dot shows the true input parameter value. The three middle panels show the distribution of maximum likelihood estimates (MLEs) for the three parameters of interest the secondary attack risk (SAR), the variation in infectiousness (p_80_), and the variation in susceptibility (s_80_). For each panel, the colored horizontal line is the actual value of the parameter used to generate the simulated data, and the white dot is the mean of the individual MLEs. The rightmost panel shows the MLE for the SAR if heterogeneities are not included in the model inference framework. Parameters were estimated for 1,750 different simulated datasets. Other model parameters were fixed as described in the Methods.

Unbiased estimates of the degree of heterogeneity in infectiousness (p_80_) or susceptibility (s_80_) can also be inferred from simulated data, albeit with significantly more uncertainty even for larger sample sizes (Figure 3A,B). For example, for simulated datasets of 5,000 individuals, a value of p_80_ = 20% could be inferred to within 18% points in 95% of cases (32% points if both heterogeneities are present) and s_80_ = 20% to within 20% in 95% of cases (34% if both heterogeneities are present).

For smaller sample sizes, the presence or absence of each type of heterogeneity can be determined reliably even if their exact value cannot be inferred. When one kind of heterogeneity was absent from simulated data (s_80_ = 80% or p_80_ = 80%) and the other was present, the best estimates for the absent heterogeneity reliably suggested that at most slight variation was present. For samples of 1,000 individuals where p_80_ = 80%, 65% of estimates for that parameter fell in the range ≤60% p_80_ ≤80%. For s_80_ = 80%, 75% of the estimates fell in that range. The maximum likelihood estimates for the parameter describing the heterogeneity that was in fact present even more reliably indicated moderate to extreme heterogeneity. For p_80_ = 20% and s_80_ = 80%, the best estimate for p_80_ was less than 40% in 94% of cases. For p_80_ = 80% and s_80_ = 20%, s_80_was less than 40% in 96% of estimates.

When both kinds of heterogeneities are present (Figure 3C), the necessary sample size for a precise measurement of all three parameters is large, upwards of thousands of households. Some of the uncertainty in the parameter governing heterogeneities in transmission dynamics comes from a degree of mutual nonidentifiability between these parameters and the SAR in smaller households (Supp Figure S1).

Overall, these analyses suggest that reasonable estimates of the amount of inter-individual variation in susceptibility and infectiousness can be made given a large enough cohort of households, and that even with smaller sample sizes the presence versus absence of these heterogeneities can be inferred.

### Quantifying heterogeneities in household studies of COVID-19

We next used our inference procedure to estimate jointly the household SAR, the variation in infectiousness (p_80_), and the variation in susceptibility (s_80_) from three different studies of COVID-19 spread in households [69, 75, 76]. We chose studies that included hundreds or more households with at least one infection, where there was likely to be a complete or high ascertainment rate of infections within the house, and with publicly-available data reporting the full distribution of household outbreak sizes (see Supplement for more details). These included a recruited cohort study by Bi et al. measuring seroprevalence in the city of Geneva, Switzerland between April and June 2020 [69]; a household contact-tracing study with PCR testing conducted by Dattner et al. in Bnei Brak, Israel between March and April 2020 with follow up serological testing conducted between May and June 2020 [75]; and an analysis of the surveillance database for PCR-positive cases in the province Ontario, Canada conducted by Tibebu et al. between July and November 2020 [76](Figure 4). The maximum likelihood estimates and 95% confidence intervals for the inferred values of each of the three transmission parameters are reported in Table 1.

**Table 1:** Estimating parameters of COVID-19 transmission heterogeneity from household studies. Estimates for household secondary attack risk † (SAR), degree of infectiousness variation (p_80_), and degree of susceptibility variation (s_80_) in three study populations (maximum likelihood estimate with 95%confidence intervals). SAR reported as a simple fraction of secondary contacts infected over total secondary contacts (discounting the follow up serological survey in the case of the results from Bnei Brak [75].) ‡ SAR reported as the best fit value from a chain binomial model with no heterogeneity.

| Population | Fit with both kinds of heterogeneity |  |  | Original report |  |
| --- | --- | --- | --- | --- | --- |
| | SAR | $p_{80}$ | $s_{80}$ | SAR | |
| Geneva, Switzerland [69] | 26% [12%, 37%] | 4% [2%, 80%] | 18% [0%, 80%] | 17%<sup>‡</sup> | [13%, 22%] |
| Bnei Brak, Israel [75] | 34% [31%, 37%] | 80% [64%, 80%] | 2% [0%, 6%] | 36%<sup>†</sup> | [34%, 38%] |
| Ontario, Canada [76] | 23% [22%, 24%] | 16% [14%, 18%] | 2% [0%, 4%] | 19%<sup>†</sup> | [19%, 20%] |

**Figure 4:**
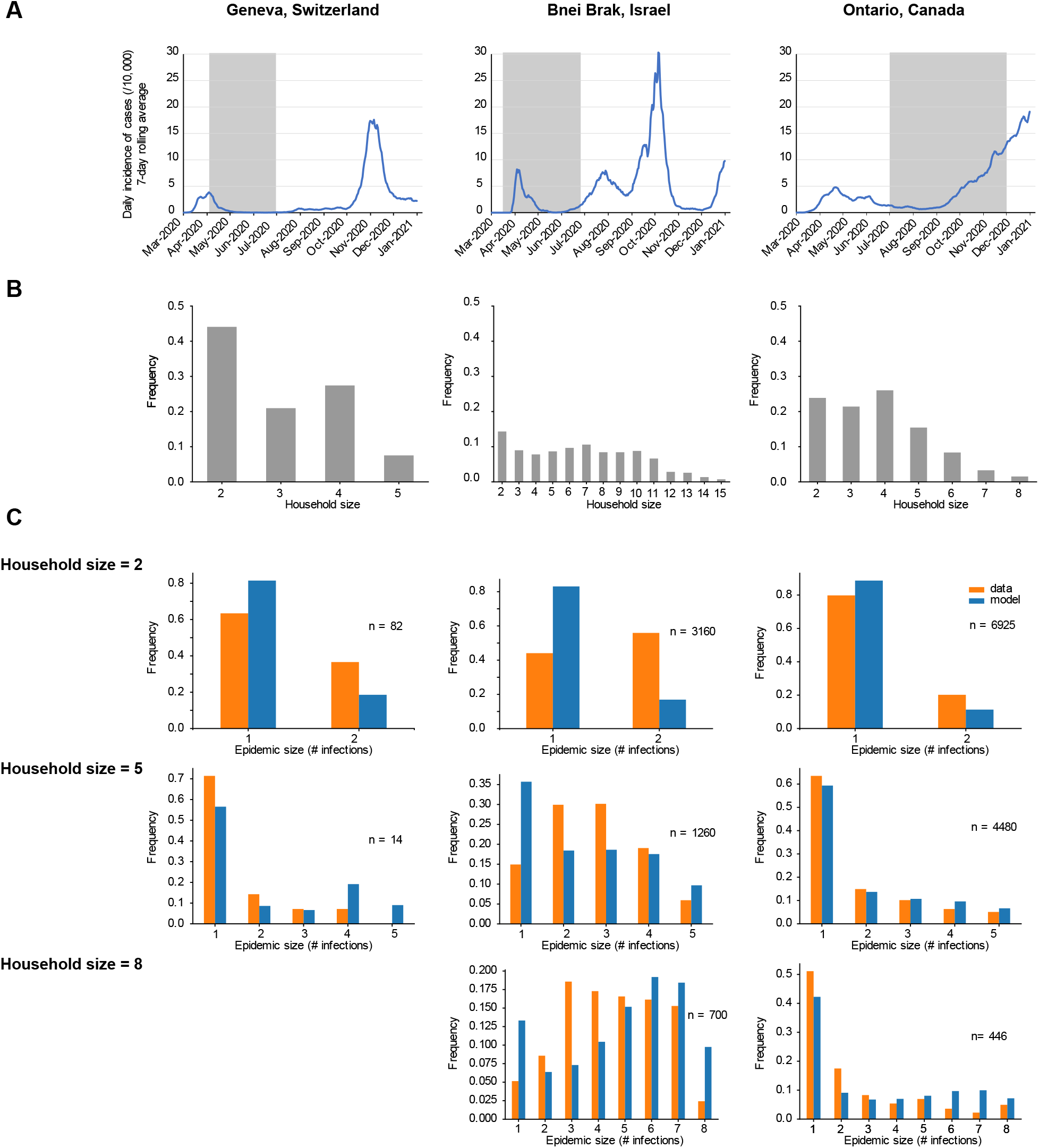
Estimating transmission heterogeneity parameters from household studies of COVID-19. (A) Timing of study (highlighted region) compared to incidence in the region throughout 2020 (daily cases per 10,000 inhabitants, 7-day rolling average) [69, 75–79]. (B) Frequency of different household sizes in the study among households with at least two members and at least one observed infection. The study by Tibebu et al. in Ontario included 358 households with more than 8 residents, but this data was not publicly available and so those households were excluded [76]. (C) Comparison of the observed distribution of household epidemic sizes (# infections) in the study (orange) with the average distribution of infections from forward simulation based on the model with the inferred maximum likelihood parameters (blue).

Of the three studies considered, the study conducted in Geneva had the most complete ascertainment of infection status but featured the fewest households. 2,267 households were enrolled, and only 181 had at least one seropositive individual with at least one secondary contact (i.e. the household size was greater than or equal to two). Households tended to be small. Among the households of interest, the average size was 3.0 and only 35% had four or more individuals (Figure 4). We estimated a household SAR of 26% (95% CI 12-37%). This was greater than the 17.3% (95% CI 13.1%-21.7%) reported in the original study though more uncertain, consistent with our results with simulated data concerning bias when heterogeneities are ignored (Figure 3). Extreme heterogeneity in both infectiousness and susceptibility was inferred from the data, though the estimates had high uncertainty, due to the lack of mutual identifiability between parameters (Supp Figure S3). This finding is also consistent with the results from testing our inference approach on simulated data, which demonstrated that a large number of households is necessary to estimate both heterogeneities precisely.

The second dataset we analyzed was from Bnei Brak, a densely populated city in Israel, and featured many large households (637 total households were considered, 62% of which had 4 or more members; Figure 4). Households entered the study when a symptomatic individual reported their illness to a healthcare provider and receiving a PCR-based diagnosis. Using our framework, we inferred an SAR of 40%, no infectiousness variation, and extreme susceptibility variation. However, we found that the best fit parameters failed to reproduce the observed pattern of infections (Figure 4C). For larger household sizes, the model overpredicts both the fraction of household outbreaks with no secondary transmission and the fraction resulting in near complete infection, whereas the data show more outbreaks of intermediate size. As such, despite the fact the best estimates for the three parameters have low uncertainty, no conclusion can be drawn about the presence or amount of heterogeneity in this population.

The largest dataset we considered came from the provincial public health surveillance database of the province of Ontario, Canada, and included 28,994 households with at least one infection and at least two residents (Figure 4B). All positive PCR tests in the province were linked back to addresses and then used to estimate the number of infections in each household. While this passive approach risks incomplete ascertainment of the infection status of individuals in a household, the widespread availability and high uptake of PCR tests in the province during the study period, especially for contacts of cases, suggest that this data may still provide a good estimate of the true household outbreak size. The maximum likelihood Absolute SAR reduction: 0.15 0.10 estimate for the household SAR was 23%. Our results also suggest the presence of substantial heterogeneity in infectiousness — with 16% of individuals being responsible for an average of 80% infections — and extreme heterogeneity in susceptibility — with 2 percent of individuals responsible for 80% of the overall susceptibility. The distribution of household infections predicted by the best fit parameters from our inference closely resembled the observed infections, suggesting our model describes the data well (Figure 4C).

### Application to the design of household intervention studies

Households are a useful setting to quantify the impact of interventions to reduce disease transmission, such as behavior change, personal protective equipment, vaccines, or treatment [60, 80, 81]. Since household cohorts are costly, it’s useful to calculate how many participants will be needed to detect the expected effect of a given intervention reliably (i.e., to “power” the study) [82]. When substantial inter-individual heterogeneity is present, there’s reason to believe that the sample size needed detect an effect will be larger than predicted based on models that don’t include heterogeneity. Using forward simulations and our inference framework, we analyzed the power of an imagined study of a household intervention that reduces the SAR between a control group (assumed SAR = 25%) and an intervention group (assumed SAR < 25%)(Table 2). Note that these power calculations concern to ability to detect the presence of an effect, not the ability to ascertain the precise SAR in either the control or intervention group.

**Table 2:**
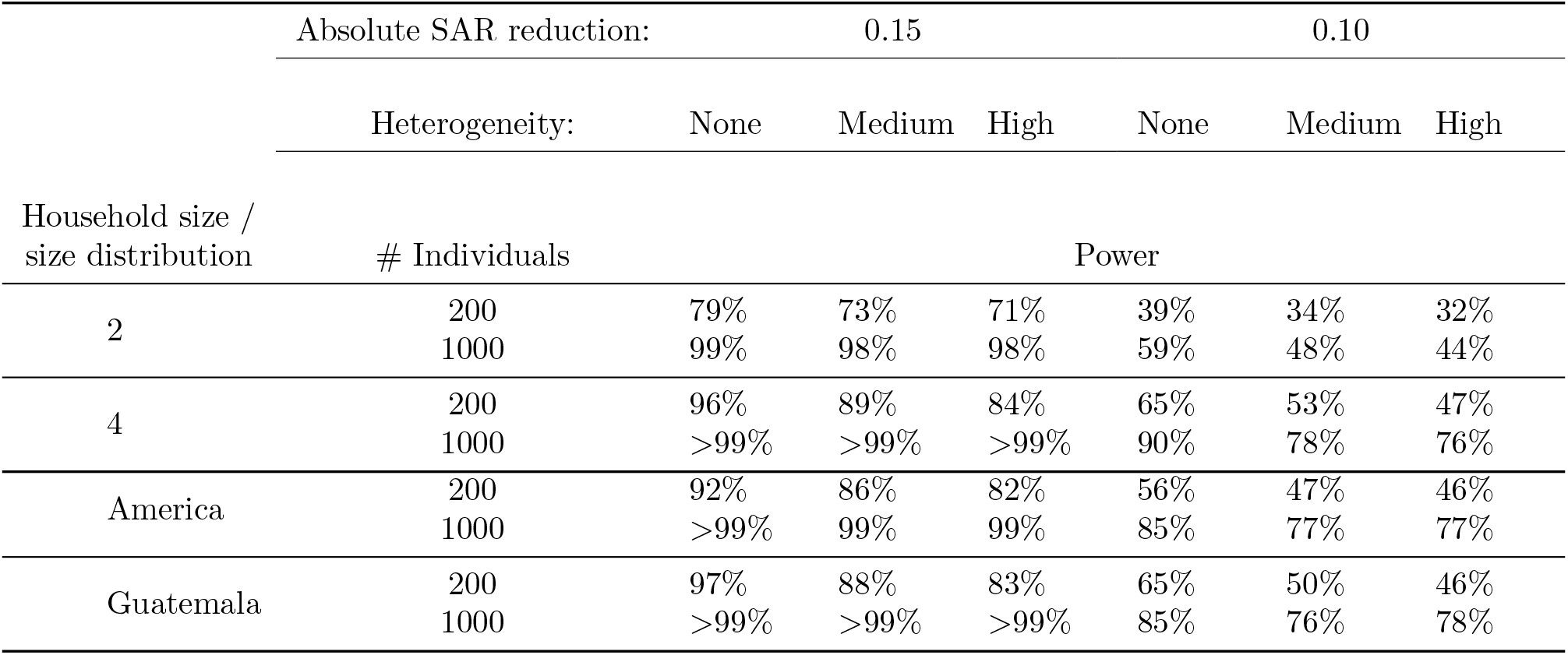
Power to measure effects of household interventions on transmission. We consider a household intervention that reduces the SAR from its baseline value in Group 1 (fixed at 25%) to a lower value in Group 2 (either 15% or 10%). 2000 simulated studies were conducted for each combination of parameters. Power is calculated using a one-sided confidence interval with significance of ff = 10%. In the scenario with “medium” heterogeneity, s_80_ = p_80_ = 50%. In the scenario with “high” heterogeneity, s_80_ = p_80_ = 20%. The distribution of household sizes for the American population was based on Census Bureau statistics [83]. For Guatemala, the approximation was based on the United Nations database of household composition [84].

In the absence of heterogeneity, we estimated that a study of 1,000 individuals divided into households of size 2 had 99% power to detect a reduction in SAR from 25% to 10% but only 59% power to detect a reduction from 25% to 15%. With a smaller study size of only 200 individuals, the powers were reduced to 79% and 39%, respectively.

We found that the amount of heterogeneity in the relative susceptibility and infectiousness of individuals in the study households had a minor and effect on the power of a study to detect a change in SAR. For the study with 200 individuals described above, the power to detect a reduction from 25% to 10% was 8% points lower (79% versus 71%) in the presence of high heterogeneity in both susceptibility and infectiousness(s_80_ = p_80_ = 20%) as compared to no heterogeneity. To detect a reduction in SAR from 25% to 15%, the power was 7% points lower (39% versus 32%). This relatively minor difference was consistent with the outcome of our experiments with simulated data, which showed that the SAR can be reliably inferred for any combination of heterogeneities present in the data-generating dynamics.

When comparing study designs with the same total number of individuals divided into households of different sizes, we found that households of size 4 produced significantly greater power for all the tested reductions in SAR and amounts of heterogeneity. For example, to detect a reduction in SAR from 25% to 15% in the absence of heterogeneity with 1,000 individuals in a study, a population divided among 250 households of size 4 provides 90% power, as compared to 59% power from the same number of individuals divided among 500 households of size 2. The relative advantage of the larger households increased as the amount of heterogeneity present increased. Larger households enjoy greater powers because the presence and effect of the heterogeneities can be more clearly separated from the role of the SAR (Figure 2). In contrast, in households of size 2, increasing s_80_ has an indistinguishable effect from decreasing SAR, which can obscure the difference in SAR between the control and the intervention arms.

Early results of our method on simulated data suggested that the precision of the MLE was determined in part by the exact distribution of sizes of the households. To determine the effect that this size distribution played in power calculations, we compared two realistic size distributions, one roughly based on American households with size greater than 1 (average size 3.0) and the other based on Guatemalan households with size greater than 1 (average size 4.4). The same power calculations performed for groups of people divided according to these distributions indicated that power increases with average household size. The powers calculated from American household sizes were between the powers for a study composed exclusively of households of size 2 and those for a study with exclusively households of size 4. The powers calculated from Guatemalan households sizes were nearly identical to powers calculated from a sample of households of size 4. For all the studies that included households of size greater than 2, there was a greater decrease in power as heterogeneity increased compared to studies with only households of size 2.

Overall, these results demonstrate that despite the need for large sample sizes to quantify the degree of infectiousness or susceptibility variation in household studies (Figure 4), more moderate study sizes can still be used to accurately quantify the SAR (Figure 4) and to estimate the effect of interventions to reduce SAR (Table 2) even in the presence of these heterogeneities.

## Discussion

We found that variation in the susceptibility and infectiousness of individuals has a significant impact on disease outbreaks within small, well-mixed populations such as households, and that the degree of this variation can be inferred from household transmission studies that report only the final outbreak size in each home. The inference method we developed combines exact forward simulation of any dynamic model – as opposed to existing methods that rely on simplified models with analytic solutions – with maximum likelihood estimation, and uses only modest and generally accessible computational resources. We showed that our inference approach can produce unbiased estimates of the household secondary attack risk (SAR), as well as the heterogeneity among individuals in both susceptibility to infection and transmissivity once infected. We found that when heterogeneity is present in the data-generating dynamics but unaccounted for in the parameter inference, estimates of the SAR become biased — especially as the amount of susceptibility variation increases. However, we identified several challenges in inferring inter-individual variability from household studies. There was high uncertainty in estimates of heterogeneity when the study population contained fewer than a few thousand individuals (highly-cited household studies of COVID-19 transmission have 500-1000 individuals [85, 86]). We observed that households of size two were generally uninformative about the heterogeneity present in susceptibility and infectiousness.

We made several important assumptions in our model. Like many other household transmission studies, we assumed that all households are well-mixed populations. More realistically, some individuals may have more intense contacts with others and secondary contacts of individuals known to be infected can take various levels of precaution to avoid infection. This contact heterogeneity could conflate our estimates of variation in infectiousness or susceptibility, meaning it is still possible that the inferred heterogeneity is caused by a mixture of behavior, biology, and host-pathogen interaction. Another key assumption was that there are no cases of multiple importations into households, meaning our results are most applicable to circumstances in which risk from the community is low and risk from household contacts is high. Lastly, we assumed that the distributions of relative susceptibility and infectiousness were agnostic to individuals’ biological traits such as age, sex, or prior immunity, and thus were identically and randomly distributed across individuals in all households. However, because our approach allows for an arbitrary model of infection spread, each of these limitations could easily be addressed in future work.

Household infection studies must make trade-offs in recruitment method, sample size, testing methodology, and index case definition because of the constraints of time, funding, and feasibility. We applied our inference procedure to three diverse studies of household spread of SARS-CoV-2, but found that only Ontario’s vast provincial database of positive PCR tests allowed us to make reliable estimates of the individual-level variation in infectiousness and susceptibility. For the Geneva-based household serosurvey [69], we could not narrow down precise estimates of heterogeneity, likely due to the small sample size. Our tests on simulated data suggested thousands of individuals were needed for precise estimation. For the Bnei Brak study [75], we found that even the best-fit model did not describe the data well, suggesting that additional sources of heterogeneity in either the infection or recruitment/testing process violated assumptions of our model. These results underscore the indispensable role that data from broad public health surveillance plays in modeling disease spread.

Estimations from the Ontario dataset suggested moderate levels of infectiousness variation and high levels of susceptibility variation, with 16% (95% CI 14-18%) of individuals responsible for 80% of secondary infections and 2% (95% CI 0-4%) of individuals responsible for 80% of population-level susceptibility. The close agreement between the distribution of household outbreak sizes in the observed data and those predicted by the best-fit model gives good reason to think that the presumed model of disease spread and individual variation described the true dynamics well. Even so, we found that a growing discrepancy between the observed and predicted number of households with many but not all individuals infected as household size increased. The passive surveillance approach used in this study is unlikely to yield full infection histories in each household, which may partially explain why our model overpredicts large household outbreaks. Further work is needed to examine the possible bias introduced by incomplete ascertainment of infection status. Contrary to the hypothesis of Tibebu et al. [76], our estimated SAR for this dataset was *higher* than the crude estimate assuming all non-index cases were infected by the primary case, even though our model allows for infections to spread over multiple generations. This fact highlights the importance of accounting for heterogeneity explicitly in order to develop a clear picture of the risk to the household contact of an infected individual.

In the case of SARS-CoV-2, age plays a role in determining susceptibility and infectiousness. In their analysis of data from Bnei Brak, Dattner et al. [75] found strong evidence that children are less susceptible than adults. The city of Bnei Brak has a young population, with 51% of individuals under the age of Our model drastically underpredicted infections in households of size 2 and overpredicted infections in large households. Declining to stratify the population by age in our inferences may have contributed to these problems with the model fit because larger households tend to contain more children and thus have a younger average age than smaller households. The enrollment process and follow up condition (return visits for testing secondary contacts were only made when a household contact reported *symptoms*) may also be the source of disagreement between data and model in this setting. Since individuals in larger households are at greater risk (due to having a large number of close contacts who are themselves at risk), under-ascertainment of infection status may censored the tail of final size distribution.

An extension of our work could take a combined approach where susceptibility varies across the population but there is a different risk factor according to an individual’s age group. Another extension could correlate susceptibility variation with household size to test the hypothesis that age primarily complicates our model by violating the assumption that individuals in households of different sizes are considered identical. Still, it’s been noted that having *a priori* knowledge of the meaningful subdivisions of a population for infection risk is especially challenging and unreliable in the study of emerging pathogens [27]. Our approach avoids the unreliability of *a priori* stratification by modeling the individual differences in a continuous — rather than a discrete — way. The high level of heterogeneity in susceptibility we inferred from COVID-19 household data highlights the utility of this approach to detect heterogeneity at the scale of a population even in the absence of known risk factors.

Our framework for estimating heterogeneity and overall rates of infection within a household is relevant not only because of what estimates imply about disease spread within a larger population, but also because of what they tell us about transmission in this vital setting for the control of epidemics. We found that even a small study can precisely estimate the SAR in the presence of heterogeneity among individuals provided that the study allows for the possibility of heterogeneity in its model of disease dynamics. Our analysis of the power of a study to detect a difference in SAR indicates that the presence of heterogeneity does not make an inference of SAR impossible. The flexibility offered by the ability to do inference with arbitrary dynamic models of infection is additionally useful because it allows our framework to be extended in the future to many kinds of household interventions such as masking or therapeutics for SARS-CoV-2 and other respiratory pathogens.

## Data Availability

All data produced are contained in the manuscript or available online at
https://github.com/tanderson11/covid_households

## Acknowledgements

This work was supported by the US National Institutes of Health grant DP5OD019851 (ALH) and R01AI146129 (MZL). We thank members of the Johns Hopkins Infectious Disease Dynamics Group and attendees of the EPIDEMICS8 meeting for helpful feedback on the project. We also thank Sarah Buchan and Semra Tibebu of Public Health Ontario for sending us the data tables used to generate their figures.

## Data accessibility

All of the code required to reproduce the results in this paper is openly available via GitHub: https://github.com/tanderson11/covid_households

