## Supplemental Methods and Figures for "Quantifying individual-level heterogeneity in infectiousness and susceptibility through household studies"

### Supplementary Methods

#### Modeling heterogeneity in infectiousness and susceptibility

##### Basic framework

Under our model, the probability that individual $j$ is infected after an exposure to an infected contact $i$ that lasts for the duration of their infection is:

$$p_{\text{infection}} = 1 - e^{-\beta\psi_j\phi_i\tau_i} \tag{S1}$$

where $\tau_i$ is the duration of the infectious period for the infected individual, $\phi_i$ is their relative infectiousness, $\psi_j$ is their contact's relative susceptibility, and $\beta$ is a model parameter that denotes the average probability per contact per unit time of transmitting infection.

The distribution of the relative infectiousness and susceptibility in the population can follow arbitrary single-parameter distributions with mean 1 and codomain $[0, \infty)$. For this paper, we model both distributions as lognormal random variables with mean 1 and variance $\text{Var}_\chi$ (where $\chi$ refers to the random variable $\phi$ or the random variable $\psi$). The parameters of the lognormal distribution — $\mu$, the mean of the log of the variable, and $\sigma$, the standard deviation of the log of the variable — are then

$$\mu_\chi = -\frac{1}{2}\log\left(\text{Var}_\chi + 1\right) \tag{S2}$$

$$\sigma_\chi^2 = \log\left(\text{Var}_\chi + 1\right) \tag{S3}$$

##### Specifying heterogeneity of infectiousness in terms of expected fraction of transmission

We quantify the heterogeneity in infectiousness in terms of the distribution of expected secondary infections. In particular, we specify the heterogeneity with one parameter, $p_{80}$, which is defined as the (smallest) fraction of individuals responsible for 80% of expected secondary infections. This value is calculated in reference to a hypothetical population that is infinite and well-mixed — not based on the expected fraction of infections caused in a population divided among households. In the limit that infectiousness across the population is constant, $p_{80} = 80\%$. As $p_{80}$ decreases, a smaller fraction of individuals is responsible for a greater fraction of infections and therefore the data is over-dispersed [74].

In order to understand how the constraint that $p_{80}$ percent of individuals are responsible for 80% of expected secondary infections fixes the distribution of relative infectiousness, we first need to calculate the expected fraction of transmission from the most infectious individuals. To individual $i$ in a population, we can assign an individual reproductive number $\nu_i = \beta\phi_i\tau_i n$ where $n$ is their hypothetical number of contacts if placed in an imagined well-mixed population. This reproductive number represents the expected secondary infections caused by that individual. Let $f_\nu$ be the probability density function (PDF) of the individual reproductive numbers of individuals. For a fixed $\nu_0$, we call the expected fraction of transmission from individuals who are expected to cause at least $\nu_0$ secondary infections $G_{\text{trans}}(f_\nu, \nu_0)$, and we calculate it as a ratio of the average secondary infections from the most infectious individuals to the average secondary infections from all individuals:

$$G_{\text{trans}}(\nu > \nu_0) = \frac{\int_{\nu_0}^{\infty} u f_\nu(u)\, du}{\int_0^{\infty} u f_\nu(u)\, du}. \tag{S4}$$

Let $f_{\text{infectiousness}}$ be the PDF of the relative infectiousness of individuals. Since the distribution of the length of the infectious period and the distribution of an individual's relative per-contact infectiousness ($\phi_i$) are independent, $f_\nu \propto f_{\text{infectiousness}}$. If we let $\phi_0 = \nu_0/\beta\tau_0 n$ be the relative infectiousness of an individual expected to cause $\nu_0$ infections, we therefore have

$$G_{\text{trans}}(\nu > \nu_0) = \frac{\int_{\phi_0}^{\infty} u f_{\text{infectiousness}}(u)\, du}{\int_0^{\infty} u f_{\text{infectiousness}}(u)\, du}. \tag{S5}$$

But the distribution of relative infectiousness $\phi_i$ is constructed so that its mean value is 1, therefore the denominator is equal to 1 and

$$G_{\text{trans}}(\nu > \nu_0) = \int_{\phi_0}^{\infty} u f_{\text{infectiousness}}(u)\, du\,. \tag{S6}$$

In order to derive from Eq. (S6) the constraint on $f_{\text{infectiousness}}$, we introduce the infectiousness $x_q$, where $q$ is any percentile, that splits the most infectious $1-q$ individuals from the least infectious $q$ individuals. Let $F_{\text{infectiousness}}$ be the cumulative distribution function of $f_{\text{infectiousness}}$ then

$$F_{\text{infectiousness}}(x_q) = 1 - q. \tag{S7}$$

Then, for a given $p_{80}$, $x_{p_{80}}$ is the infectiousness that divides the most infectious $p_{80}$ percent of individuals from the rest. As a result, our way of quantifying heterogeneity in relative infectiousness in terms of $p_{80}$ boils down to the following two constraining equations:

$$F_{\text{infectiousness}}(x_{p_{80}}) = 1 - p_{80} \tag{S8}$$

$$0.8 = \int_{x_{p_{80}}}^{\infty} u f_{\text{infectiousness}}(u)\, du\,. \tag{S9}$$

For our specific choice that relative infectiousness follows a lognormal distribution then $f_{\text{infectiousness}}$ and $F_{\text{infectiousness}}$ are the PDF and CDF of $\text{Lognormal}(-\frac{1}{2}\log(\text{Var}_\phi + 1), \log(\text{Var}_\phi + 1))$ per Eqs.(S2) and (S3). We calculate the variance $\text{Var}_\phi$ of the lognormal random variable of relative infectiousness by simultaneously solving Eq. (S8) and Eq. (S9) using `scipy` least squares optimization.

##### Specifying susceptibilities through $s_{80}$

As an infection spreads through a population with heterogeneous susceptibilities, the average susceptibility in the uninfected population changes over time. On average, the more susceptible individuals are infected first, which causes the average susceptibility in the remaining fraction of the population to decrease over time.

We specify the distribution of susceptibilities through the quantity $s_{80}$, which we define as the smallest fraction of the population that must be infected in order to reduce the total remaining susceptibility to 80% of its initial total. When there is no variation in the individual susceptibilities, $s_{80} = 0.8$. If $s_{80} = 0.2$, then the most susceptible 20% of individuals are responsible for 80% of the total susceptibility.

As before for infectiousness, let $f_{\text{susceptibility}}$ and $F_{\text{susceptibility}}$ be the PDF and CDF respectively of the distribution of relative per-contact susceptibility in the population. For a fixed value of $s_{80}$, let $y_{s_{80}}$ be the corresponding susceptibility that divides the top $s_{80}$ percent of individuals from the bottom $1 - s_{80}$ percent such that $F_{\text{susceptibility}}(y_{s_{80}}) = 1 - s_{80}$. Then:

$$0.8 = 1 - \int_{0}^{y_{80}} u f_{\text{susceptibility}}(u)\, du\,. \tag{S10}$$

Note that the math here is identical to the math used to parametrize the distribution of individual infectiousness. In other words, if $s_{80} = p_{80}$, then the variance of the distribution of infectiousness is equal to the variance of the distribution of susceptibility.

##### Relating a specified secondary attack risk to the underlying transmission probability $\beta$

The model parameter $\beta$ quantifies the probability per unit time that infection spreads between an average infectious individual (with relative infectiousness 1) an average susceptible contact (with relative susceptibility 1). In the absence of heterogeneity or variable infection lengths, $\beta$'s value is defined by the relationship $p_{\text{escape}} = e^{-\beta\tau}$.

While performing inferences, we choose to specify the household secondary attack risk as a model input and derive the value of $\beta$ that achieves that household secondary attack risk (SAR). We define the SAR as the probability of infection per household contact caused by a single initial introduction of the pathogen (i.e., not taking into account chains of infection beyond the initial → secondary). SAR can be calculated as follows:

$$\text{SAR} = 1 - \langle p_{\text{escape}} \rangle. \tag{S11}$$

We choose to perform inferences with respect to SAR because it is often the quantity of interest in empirical studies. SAR can be roughly estimated using statistical averages without the aid of a dynamic model or forward simulation, and it conveys relevant public health information: the risk to secondary contacts from a single infection in a household.

Let $H = \tau\phi\psi$ be a random variable that is the product of the random variables that describe the distribution of infection periods $\tau$, the distribution of infectiousness $\phi$, and the distribution of susceptibility $\psi$, and let $h$ be its PDF. In other words, $H$ is the effective period of infection. By analogy to the relationship $p_{\text{escape}} = e^{-\beta\tau}$, we have

$$\text{SAR} = 1 - \int_0^\infty e^{-\beta x} h(x)\, dx\,. \tag{S12}$$

Because we use lognormal random variables to describe individual susceptibility, individual infectiousness, and the duration of the infectious state, and the product of lognormal random variables is another lognormal random variable, it follows that $H = \text{Lognormal}(\mu_\tau + \mu_\psi + \mu_\phi, \sigma_\tau^2 + \sigma_\psi^2 + \sigma_\phi^2)$.

We specify SAR and the distributions of state lengths, infectivities, and susceptibilities, and we use them to calculate the model parameter $\beta$. We can evaluate the integral in question numerically in order to solve for $\beta$ that produces the desired SAR.

#### Computational methods

Stochastic forward simulation was conducted in each household by performing an exact forward simulation based on the Gillespie method [88], modified to allow the duration of the exposed and infectious states to follow an arbitrary distribution (rather than only an exponential distribution). The code was written in Python and used the 'torch' package to implement fast calculations on the GPU and to sample pseudorandom numbers.

For each household size, simulation was conducted at each point in a three-dimensional grid of parameters. This regular grid was composed of all combinations of three parameters from the following ranges:

$$\begin{aligned} s_{80} &\in [0.02, 0.04, 0.06, \ldots, 0.80] (40 \text{ values total}) \\ p_{80} &\in [0.02, 0.04, 0.06, \ldots, 0.80] (40 \text{ values total}) \\ \text{SAR} &\in [0.01, 0.02, \ldots, 0.60] (60 \text{ values total}). \end{aligned}$$

At all points, we simulated 200,000 households of each size. This data was used in assessing the likelihood of parameter values (and therefore the maximum likelihood estimate) for simulated data (Figure 3) for empirical data (Figure 4 and Table 1) and for power calculations for household interventions (Table 2)

#### Dependence of reliability of inference on the distribution of household sizes

We noted that reliable inference of $s_{80}$, $p_{80}$ and SAR is possible with a sufficient sample size (Figure 3). This result was based on simulated data generated by simulating household infections for individuals divided among households of size $\geq 2$ is accordance with data from the US census (average size of 3.0 individuals).

We repeated this analysis for a variety of different household size distributions (Supp Figure S1), and we found that the precision and reliability of the fit generally increases with a larger average household size. Estimates of SAR are not particularly sensitive to this effect, consistent with our result that SAR estimates are precise for a wide range of sample sizes. For estimates of $s_{80}$ there is a pronounced effect where larger average household sizes correspond to much better estimates of the amount of susceptibility variation (regardless of the kinds and amount of heterogeneity present in the data-generating dynamics). For estimates of $p_{80}$ the distribution of individuals among households of size 3,4, and 5 (average size: 3.83) produced the most precise estimates, with the populations with smaller and larger average household sizes both yielding a greater amount of uncertainty in the estimates of $p_{80}$.

For the population based on American households, we found bias in the estimates of SAR for a model that discounts heterogeneity only when there was substantial variation in susceptibility (Figure 3A). When considering the affect of average household size, we found that a larger average household size produces an increasing bias in the the estimate for SAR when variation in infectiousness is present but discounted by the model used for inferring SAR (Supp Figure S1 top right). For the simulated population with average size 2.77, the estimate for a SAR with a misspecified model was centered on 19%. For the population with average size 3.83, the estimate was centered on 18%. For the population with average size 4.86, the estimate was centered on 17%. This bias increases with average household size because the infection risk to each secondary contact of an infected individual is correlated with that individual's infectiousness. This behavior causes the misspecified model to depart more from the actual dynamics when infectiousness varies as household size increases.

#### COVID-19 household data

Numerous household studies of the spread of COVID-19 have been conducted since the pandemic first spread across the globe in 2020 [89–91]. Three major sets of factors distinguish different studies. The first set is the content of the sample — including the methodology for entering individuals into the study, the number of individuals enrolled, and how those individuals are divided among households — which plays a role in determining possible sources of error, factors that may need to be controlled for, and what inferences are possible. The second set is the timing with respect to the waves of epidemic: the probability of importation (and subsequent multiple importation) depends on the overall level of spread in the population and affects the validity of a model that assumes there is a single index case within each household [92]. The third set is the means of ascertaining the infection status of individuals and whether that test was applied to all individuals, only symptomatic individuals, or a mix of symptomatic individuals and consenting asymptomatic individuals. Most household studies of COVID-19 ascertain the infection status of individuals by testing all contacts regardless of symptoms via reverse transcription polymerase chain reaction (RT-PCR) at least once and typically twice or more [90].

Based on our model, an ideal study would have a large sample of households with three or more individuals, be conducted during a period of low community risk, and enter households into the study randomly to avoid observation bias of symptomatic individuals, and fully ascertain the seropositivity of individuals regardless of symptoms. Due to many practical constraints, such studies are rare. We reviewed approximately twenty-five household studies and selected three studies for further analysis based on their diverse set of characteristics and their efforts to approximate these ideal circumstances. We located studies by searching for key terms such as "household transmission," "attack rate," and "secondary infections" between June and September of 2021. For the majority of studies that we excluded, we excluded them on the grounds that they had fewer than two hundred total positive cases, which initial theoretical investigations revealed to be insufficient to make precise estimates of our three parameters of interest. Another discriminant among studies was the availability of data. Some studies that relied on passive surveillance databases, such as the UK's HOSTED database did not provide open access to their data tabulated by household size and were therefore unsuitable for our purposes. We preferred studies of households from the initial wave of the pandemic in 2020, as such studies avoided the complications of prior immunity and variant emergence. Since no perfect studies for our purposes existed, we made compromises that prioritized having a diverse range of studies included in our analysis in hopes of gaining more insight into what qualities of study made it tractable for our analysis.

##### Methodological notes for particular studies

The study in the city of Bnei Brak, Israel, initially ascertained infection status by RT-PCR testing of symptomatic individuals and consenting asymptomatic individuals but conducted a follow up serostudy to estimate the false negative rate [75]. We applied a randomized correction to the reported number of infections in each household by generating 20 studies where individuals reported as negative were considered positive at random with probability given by the reported correction factor of the study authors.

The study in the province of Ontario, Canada included 358 households (1.2% of all households) with 9 or more individuals. These households were pooled in the reported data in such a way that a household of size 9 with 4 secondary infections (4/8 secondary contacts infected) was indistinguishable from a household of size 13 with 6 infections. We excluded this small fraction of households and restricted our analysis to households with between 2 and 8 individuals.

##### Contours of likelihood for household studies

We used our framework to infer SAR, $s_{80}$, and $p_{80}$ in the context of three household studies: one conducted in Bnei Brak, Israel; one conducted in Geneva, Switzerland; and one conducted in Ontario, Canada. In addition to calculating a maximum likelihood estimate (MLE) and 95% credible intervals, we also calculated a three-dimensional likelihood surface (equivalently, a probability density of the posterior with uniform priors) for those parameters.

This surface can encode valuable insights even when a precise estimate of one or more parameters is not possible. For the study conducted in Geneva, the 95% confidence intervals for the estimates $s_{80}$ and $p_{80}$ showed extremely high uncertainty in the MLE for these parameters. The contours of the likelihood surface (Supp Figure S3) indicate that this uncertainty is due to a steep trade-off between the parameters that govern heterogeneity ($s_{80}$ and $p_{80}$) and the parameter that governs average risk to secondary contacts SAR.

For the studies conducted in Bnei Brak and in Ontario, the probability density was localized almost entirely to a single point on the discrete grid of points where the likelihood was calculated (Supp Figure S2 and Supp Figure S4). The lack of trade offs between the parameters and the lack of uncertainty is due to two factors. First and most importantly, these two studies featured many times more households (Figure 4C) than the study in Geneva. More households in the sample steepens the multinomial likelihood (Equation 1): an equal deviation between the observed and expected frequencies of infection for a given set of parameters $\theta$ produces a lower likelihood if there are more households included the observation. Second, these two studies also featured more many-person households than the study in Geneva, which is known to correspond to greater certainty in the estimates of parameter values (Figure 3). That both the Bnei Brak study and the Ontario study produced localized contours but the model of best fit for Bnei Brak failed to reproduce the observed infections underscores the importance of judging the fit of a model in absolute terms and not only based on the narrowness of the 95% CI.

### Supplementary Figures

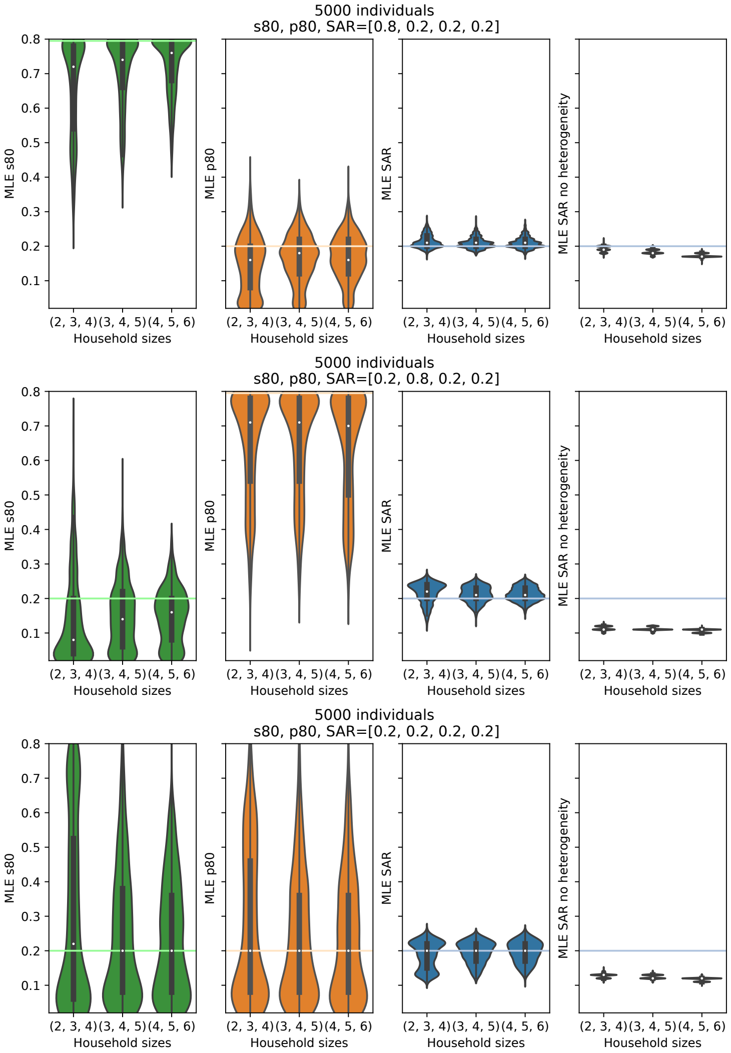


Figure S1: **Maximum likelihood estimates for SAR and degree of heterogeneity on simulated data based on the sizes of included households.** Simulated datasets were generated using 5,000 individuals divided evenly among households of three different sizes. In each plot, the leftmost violin represnts the distribution of predictions when individuals are divided evenly among households of size 2, 3, and 4 (average size: 2.77). The center: 3, 4, and 5 (average size: 3.83). The right: 4, 5, and 6 (average size 4.86). A) Simulated data includes only variation in infectiousness ($p_{80}$). B) Simulated data includes only variation in susceptibility ($s_{80}$). C) Simulated data includes variation in both infectiousness and susceptibility. The three left panels show the distribution of maximum likelihood estimates (MLEs) for the three parameters of interest - the secondary attack risk (SAR), the variation in infectiousness ($p_{80}$), and the variation in susceptibility ($s_{80}$). For each panel, the colored horizontal line is the actual value of the parameter used to generate the simulated data, and the white dot is the mean of the individual MLEs. The rightmost panel shows the MLE for the SAR if heterogeneities are not included in the model inference framework. Parameters were estimated for 1,500 different simulated datasets. Other model parameters were fixed as described in the Methods.

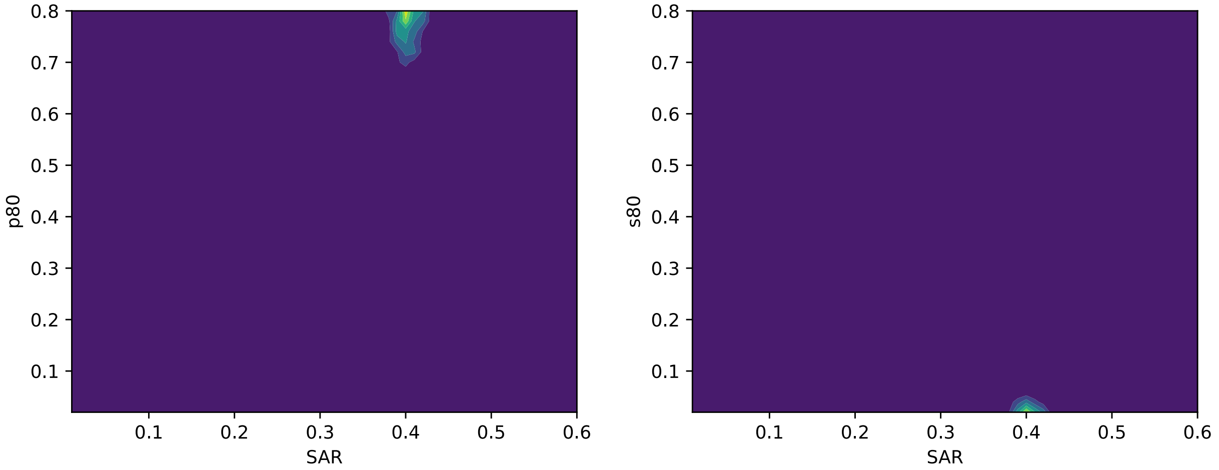


Figure S2: Likelihood surfaces describing the probability of the model parameters given the observation of the study conducted in Bnei Brak, Israel by Dattner et al. [75]. Yellow is more likely, blue is less likely. Left: likelihood plotted as a function of SAR and $p_{80}$. Right: likelihood plotted as a function of SAR and $s_{80}$. Each two dimensional surface marginalizes over the third, not shown, parameter.

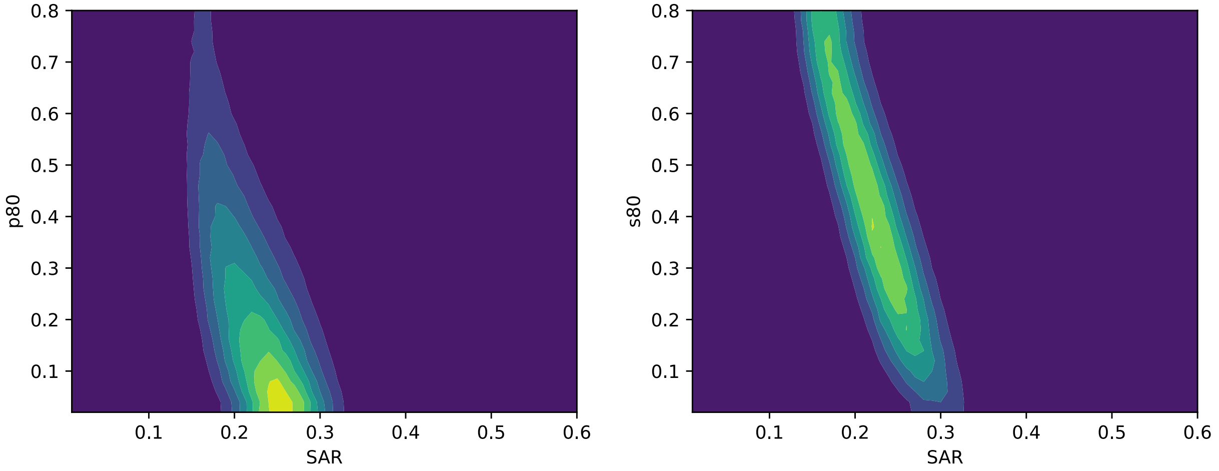


Figure S3: Likelihood surfaces describing the probability of the model parameters given the observation of the study conducted in Geneva, Switzerland by Bi et al. [69]. Yellow is more likely, blue is less likely. Left: likelihood plotted as a function of SAR and $p_{80}$. Right: likelihood plotted as a function of SAR and $s_{80}$. Each two dimensional surface marginalizes over the third, not shown, parameter.

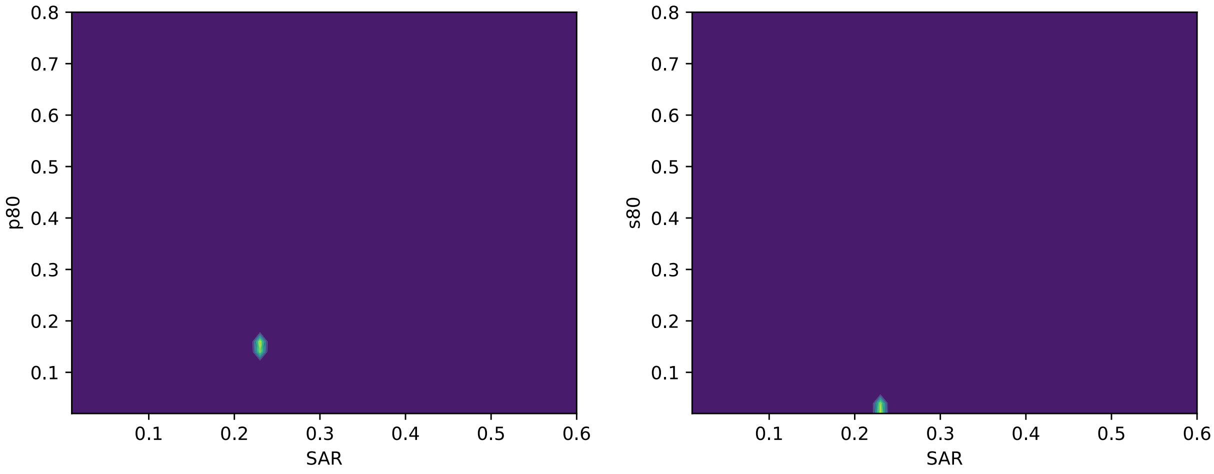


Figure S4: Likelihood surfaces describing the probability of the model parameters given the observation of the study conducted in Ontario, Canada by Tibebu et al. [76]. Yellow is more likely, blue is less likely. Left: likelihood plotted as a function of SAR and $p_{80}$. Right: likelihood plotted as a function of SAR and $s_{80}$. Each two dimensional surface marginalizes over the third, not shown, parameter.
